# Investigating the oxidative stress-vascular brain injury axis in mild cognitive impairment of the Alzheimer type

**DOI:** 10.1101/2024.09.24.24313962

**Authors:** Flavie E. Detcheverry, Sneha Senthil, Winnie L.K. Motue, Chris Hosein, Rozie Arnaoutelis, David Araujo, Dumitru Fetco, Haz-Edine Assemlal, Samson Antel, Douglas L. Arnold, Jamie Near, Hyman M. Schipper, AmanPreet Badhwar, Sridar Narayanan

## Abstract

**INTRODUCTION:** Oxidative stress may contribute to brain injury in Alzheimer disease (AD) continuum. The antioxidant glutathione (GSH) can be assessed with magnetic resonance spectroscopy (MRS). Since the relationship between GSH and vascular-brain injury is unknown in the AD continuum, we address this gap in mild cognitive impairment (MCI).

**METHODS:** 3T MRI/MRS data were obtained from 31 MCI participants. GSH and total N-acetylaspartate (tNAA; neuroaxonal integrity marker) were measured in posterior cingulate cortex (PCC) and frontal white matter (FWM). Cerebrovascular injury was assessed using white matter hyperintensity (WMH) volume. Global and regional brain tissue integrity were assessed using normalized brain (NBV) and hippocampal volumes.

**RESULTS:** Significant associations were reported in FWM between GSH/total creatine (tCr) and tNAA/tCr, and between GSH and both WMH and NBV. tNAA, GSH/tCr, and tNAA/tCr were higher in PCC than FWM.

**DISCUSSION:** Our results suggest that oxidative stress contributes to vascular-brain injury in MCI.

## 1. INTRODUCTION

Age-related dementias impact over 44 million people worldwide, a number projected to triple by 2050 as the global population ages [1]. Alzheimer’s disease (AD), the most prevalent of these dementias [2], is often preceded by a prodromal stage known as mild cognitive impairment (MCI) [3]. The underlying pathology of AD is a complex, multifactorial process that begins decades before dementia onset, typically during midlife [4–6]. Traditionally, AD progression has been characterized by the sequential accumulation of amyloid-beta plaques, tau tangles, and brain atrophy [4–6]. However, recent updates to the AD biomarker system acknowledge the multifaceted nature of the disease by incorporating additional factors such as inflammation and vascular brain injury, both of which play crucial roles early in AD progression [7–10]. Oxidative stress has emerged as a major driver and consequence of both inflammation and vascular injury in AD [11–14], prompting clinical trials of antioxidant therapies as potential disease-modifying treatments [15].

Oxidative stress arises when the delicate balance between reactive oxygen species (ROS) and antioxidants is disrupted, either due to increased ROS production or weakened antioxidant defences [16–18]. In AD, substantial evidence points to persistent oxidative stress within brain tissue throughout the disease course [11]. Amyloid-beta oligomers are believed to contribute significantly to this process, initiating oxidative stress in the early stages of the disease [19]. This imbalance between ROS and antioxidants damages nucleic acids, lipids, proteins, and other molecules, triggering inflammation, cell death, and tissue damage [18,20]. Glutathione (GSH), the brain’s most abundant antioxidant, is crucial in defending against ROS-induced damage [17,18]. It acts by either directly neutralizing ROS or participating in enzymatic reactions [18]. GSH levels in humans can be monitored using magnetic resonance spectroscopy (MRS), a powerful non-invasive method for assessing cerebral metabolites, or through biochemical assays of brain tissue and biofluids (e.g., blood) [18].

Although research is still ongoing, most studies report perturbed GSH levels in brain and peripheral biofluids in the MCI and dementia stages of the AD continuum compared to healthy controls [21–28]. In particular, *in vivo* 3T MRS studies using edited sequences have revealed lower GSH levels in key brain regions in AD dementia individuals [21–24]. While reductions in GSH, specifically in the hippocampus and frontal cortex, have been linked to poorer cognition [22], this association has not yet been explored across all brain regions. Lower GSH levels have also been detected in the blood of AD dementia individuals [26]. This widespread decline in GSH further reinforces the link between oxidative stress and AD pathology [22]. Supporting this, longitudinal data indicate that higher baseline plasma GSH levels in cognitively unimpaired older adults are associated with a lower risk of developing AD and better long-term preservation of executive function [29].

Despite some understanding of GSH levels and their connection to cognition in AD, the relationship between GSH and both neuronal and vascular brain injury in AD remains poorly defined. While lower GSH levels are associated with increased white matter hyperintensity (WMH) volume, a marker of vascular brain injury, in non-AD conditions [30], this association in AD has yet to be explored. This knowledge gap is critical given the role of oxidative stress in both vascular dysfunction and neurodegeneration. Our study aims to address this gap by focusing on individuals with MCI of the AD type, investigating how brain GSH levels relate to markers of (a) vascular brain injury, (b) neuroaxonal damage, and (c) overall brain integrity. Additionally, we examine the association between a cognitive screening test, the Montreal Cognitive Assessment (MoCA) [31], and vascular injury markers in MCI, aiming to clarify the interplay between oxidative stress, vascular pathology, and cognitive decline.

## 2. METHODS

### 2.1 Participants

Thirty-one MCI participants (20 females) were recruited from the Jewish General Hospital Memory Clinic for an open-label, single-arm pilot study testing the ability of a whey protein dietary supplement, a natural source of the glutathione precursor cysteine, to increase brain GSH in people with MCI (NCT03448055). The work reported here employs the <u>baseline data</u> from that study. The MCI diagnosis was based on clinical assessment, which comprised detailed neuropsychological testing (including MoCA), blood work, imaging (computed tomography and/or MRI) and family, personal, and medical history, according to formal criteria of McKhann et al. [32] and the ICD-10. The study was approved by the research ethics boards at the CIUSSS-Centre-Ouest de Montreal for the Jewish General Hospital and the McGill University Health Centre for the Montreal Neurological Institute-Hospital, and informed consent was obtained for all participants.

### 2.2 Brain MRI/MRS data acquisition

All participants were scanned using a 3 tesla (T) Siemens Prisma*fit* MR scanner (Siemens, Erlangen, Germany) at the McConnell Brain Imaging Centre of the Montreal Neurological Institute-Hospital using a body coil transmitter and a 64-channel head receive array coil. The MRI protocol included a whole brain, 3D T1-weighted (w) magnetization-prepared-2 rapid acquisition of gradient echos (MP2RAGE) sequence (1 mm^3^ isotropic resolution) for structure segmentation and a 3D T2w fluid-attenuated inversion recovery (FLAIR) sequence (1 mm^3^ isotropic resolution) for WMH segmentation. The MRS protocol included a single voxel SPin Echo full Intensity Acquired Localized (SPECIAL) sequence [33] in two brain regions relevant to MCI pathology of the AD-type, the posterior cingulate cortex (PCC [34]) and left frontal white matter (FWM [22]), with the following scan parameters: repetition time (TR)/echo time (TE) = 3000/8.5 ms, 128 averages (PCC; Figure 1A) and 256 averages (FWM; Figure 1B), 2048 acquired sample points, VAPOR (variable power and optimized relaxation delays) water suppression [35], 2000 Hz spectral width and delta frequency = −2.3 ppm. For each MRS voxel, 16 averages of water unsuppressed data were also acquired for eddy-current correction and water referencing. Six outer volume suppression (OVS) slabs were manually placed around the volume of interest (VOI) to suppress unwanted lipid signals. Shimming was performed automatically using the Siemens “brain” B0 shim mode. The total MR acquisition time was about 60 min.

**Figure 1.**
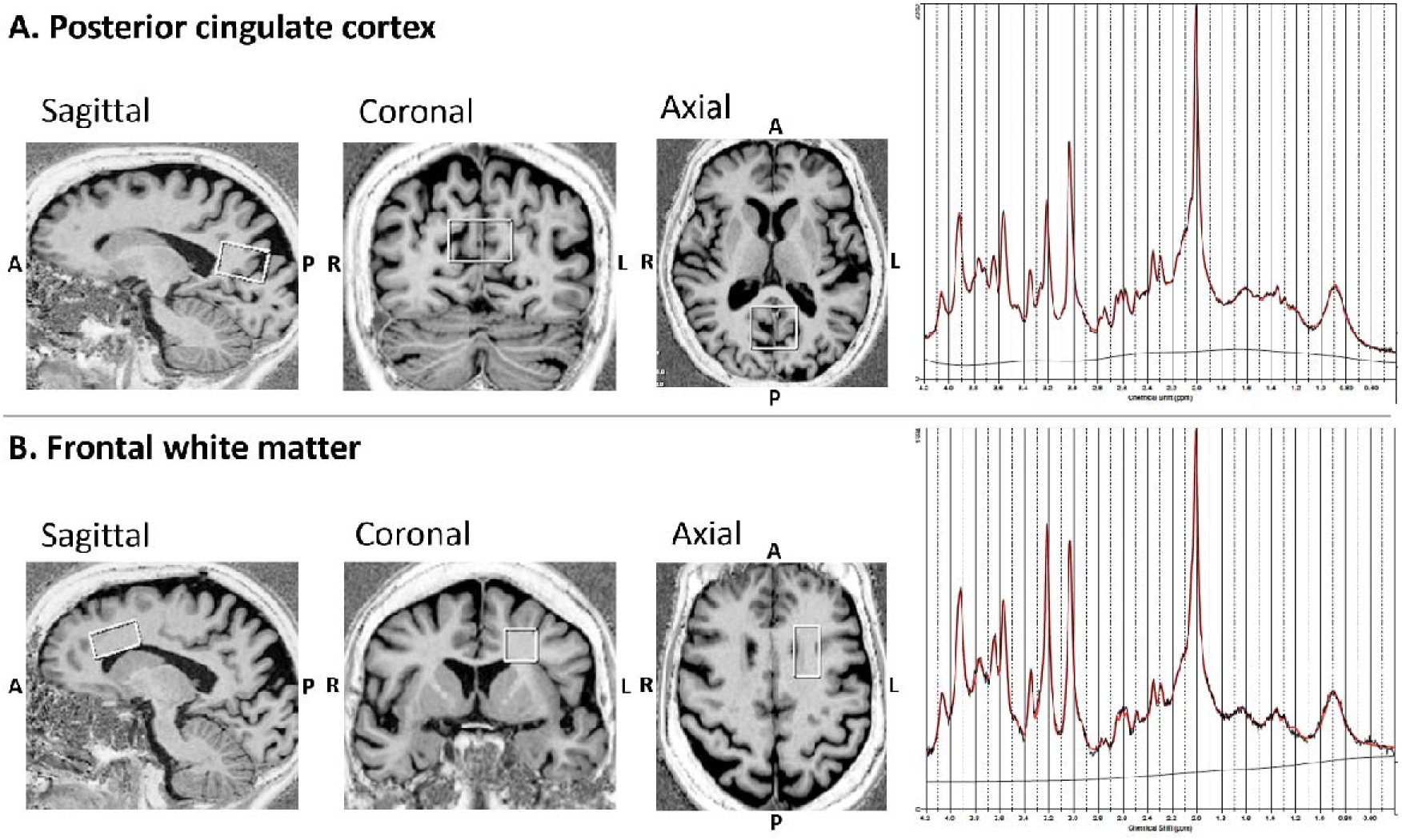
MRS voxels in **A)** posterior cingulate cortex (3 x 3 x 2 (RL x AP x FH) cm³), and **B)** frontal white matter (1.5 x 3 x 1.5 (RL x AP x FH) cm³). MRS spectra from the respective regions are displayed in the right panel. *Abbreviations:* MRS, magnetic resonance spectroscopy.

### 2.3 Data preprocessing

The acquired MRS data were exported from the scanner console in TWIX (.dat) format. The data was preprocessed using the FID-A toolkit (github.com/CIC-methods/FID-A). The pipeline performed the following series of preprocessing steps: (1) weighted coil combination [36]; (2) retrospective frequency and phase drift correction of the resulting spectral averages using spectral registration [37]; (3) Removal of motion-induced bad averages; and (4) signal averaging.

### 2.4 MRS quantification

Metabolite quantification was performed using the LCModel software (Stephen Provencher, Inc., Oakville, Canada) [38]. The water unsuppressed scan was used for internal referencing and eddy current correction. The basis set was generated in FID-A, consisting of the following 19 brain metabolites: Alanine (Ala), Aspartate (Asp), Phosphocholine (PCh), Creatine (Cr), Phosphocreatine (PCr), gamma-aminobutyric acid (GABA), glutamine (Gln), glutathione (GSH), glycine (Gly), myo-inositol (Ins), N-acetylaspartate (NAA), scyllo-inositol (Scyllo), taurine (Tau), glucose (Glc), N-acetylaspartyl-glutamate (NAAG), glycero-phosphocholine (GPC), phosphatidylethanolamine (PE), serotonin (Ser) and ascorbic acid (Asc). Basis spectra for 9 macromolecules (MM) were simulated and incorporated in the basis set. Specifically, the frequencies were: 0.89, 1.2, 1.39, 1.63, 1.98, 2.28, 2.98, 3.19, and 3.75 ppm. The LCModel default values for WCONC, ATTH2O, and ATTMET were used, and additional correction for T1-relaxation or T2-relaxation of water and metabolites was not performed, given the relatively long TR and very short TE. Spectra were fit between the range [0.2 - 4.2 ppm]. The metabolites of interest for this study were the oxidative stress marker GSH and the neuroaxonal integrity marker tNAA (NAA+NAAG).

Further details of the MRS acquisition and analysis are also included in the MRS reporting checklist (Supplementary Table S1).

### 2.5 Vascular brain injury measurements

WMHs were segmented in both supratentorial and infratentorial brain regions by a locally developed automated, multispectral Bayesian technique using coregistered FLAIR and MP2RAGE T1w images. The automated WMH masks were reviewed and manually corrected by an experienced medical image analyst, using the DISPLAY software package (https://www.bic.mni.mcgill.ca/software/Display/Display.html). WMH volume was then calculated from the corrected masks. In addition, a qualitative assessment of WMH severity was performed by an experienced neuroradiologist using the Fazekas scale, with a score of 0 reflecting the absence of WMH, a score of 3 reflecting a significant WMH burden characterized by confluent white matter lesions, and 1 denoting an intermediate burden [39].

### 2.6 Brain volume measurement

Brain volume was segmented on the T1w MP2RAGE images and estimated using the locally-developed Brain Tissue Composition-Net (BTCNet) pipeline, based on a published framework [40]. This pipeline involves generating tissue segmentation masks using a custom 3D CNN, inspired by the U-Net architecture. The model was trained on an internal dataset of co-registered T1w MRI scans with 1 x 1 x 1 mm^3^ isotropic sampling, comprising 130 participant sessions. The ground truth for healthy tissue segmentation was established using an automated multi-atlas label fusion method [41]. The native output was multiplied by a scaling factor to yield the head-size normalized brain volume [42]. Quality control was performed by visual inspection.

### 2.7 Hippocampal volume measurement

The total hippocampal volumes were segmented on T1w MP2RAGE images using an in-house hippocampal segmentation pipeline that consisted of three main parts described in previous studies: (1) generation of a patient template image [43], (2) patch-based segmentation [41], and (3) referencing of the hippocampal template based on a harmonized protocol [44]. The native hippocampal volumes were scaled to obtain the head-size normalized hippocampal volumes, using a scale factor derived from a skull-based registration of each individual’s T1w MRI to the MNI-ICBM152-2009c (https://nist.mni.mcgill.ca/atlases/) standard space template [42,43]. To ensure accuracy of measurements and to identify any potential issues in the segmentation or scaling process, a visual quality inspection was performed by a neuroradiologist.

### 2.8 Statistical analysis

All statistical analyses were performed using Python (version 3.9.13). Relationships between GSH levels and markers of vascular brain injury, namely, WMH volume, Fazekas score; the neuroaxonal integrity marker tNAA; markers of normalized brain and hippocampal volume; and cognition assessed using the MoCA, were assessed using Pearson’s correlation. A paired two-tailed *t*-test was performed between ROIs for a given metabolite, corrected for multiple comparisons (FDR = 5%). The significance threshold for all statistical tests was set at *p* < 0.05.

## 3. RESULTS

### 3.1 Demographics

A total of thirty-one MCI participants (age range: 55-86 years of age) were enrolled in the study, including 20 women (Table 1). Sixteen participants had vascular risk factors and/or comorbidities, including hypertension (N = 12), dyslipidemia (N = 8), and coronary artery disease (N = 3). The mean MoCA score was 24.68 (range: 20-29). The mean Fazekas score was 0.74 (range: 0-2). Age was correlated (a) positively with Fazekas scores and WMH volume, and (b) negatively with GSH levels in the PCC and hippocampal volume.

**Table 1.**
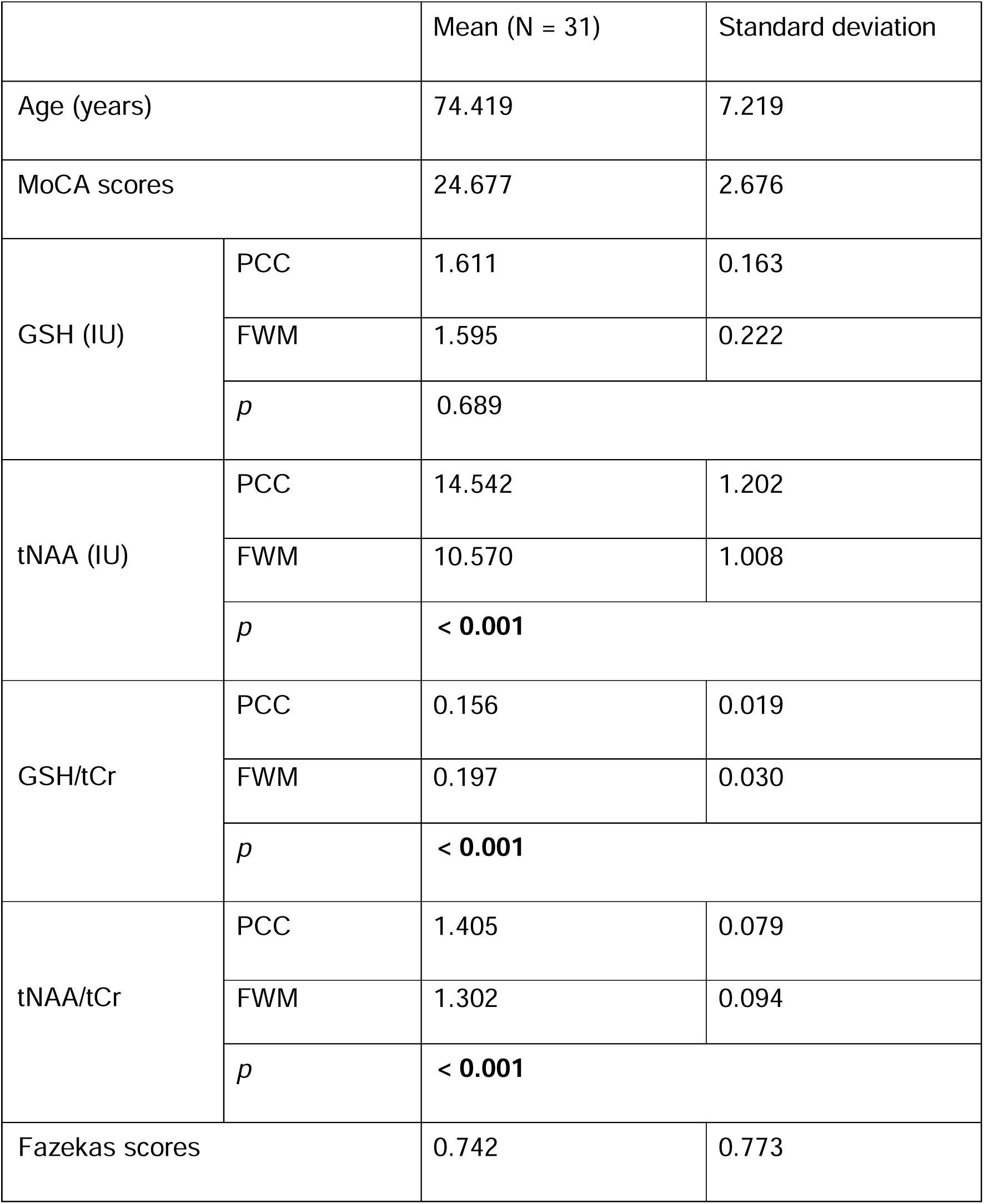
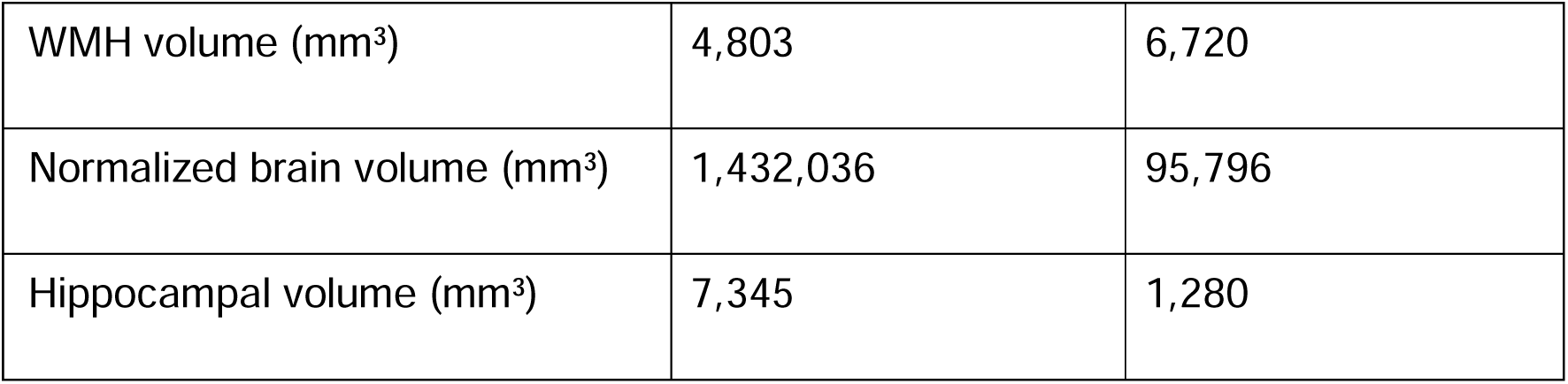
*Abbreviations:* GSH, glutathione; MoCA, Montreal cognitive assessment; tCr, total creatine; tNAA, total N-acetyl aspartate; IU, institutional units.

|  |  | Mean (N = 31) | Standard deviation |
| --- | --- | --- | --- |
| Age (years) |  | 74.419 | 7.219 |
| MoCA scores |  | 24.677 | 2.676 |
| GSH (IU) | PCC | 1.611 | 0.163 |
|  | FWM | 1.595 | 0.222 |
|  | *p* | 0.689 |  |
| tNAA (IU) | PCC | 14.542 | 1.202 |
|  | FWM | 10.570 | 1.008 |
|  | *p* | **< 0.001** |  |
| GSH/tCr | PCC | 0.156 | 0.019 |
|  | FWM | 0.197 | 0.030 |
|  | *p* | **< 0.001** |  |
| tNAA/tCr | PCC | 1.405 | 0.079 |
|  | FWM | 1.302 | 0.094 |
|  | *p* | **< 0.001** |  |
| Fazekas scores |  | 0.742 | 0.773 |

**Table 1.**
|  |  |  |
| --- | --- | --- |
| WMH volume (mm<sup>3</sup>) | 4,803 | 6,720 |
| Normalized brain volume (mm<sup>3</sup>) | 1,432,036 | 95,796 |
| Hippocampal volume (mm<sup>3</sup>) | 7,345 | 1,280 |

### 3.2 Association between oxidative stress marker and brain vascular injury marker

We found GSH levels to be negatively correlated with WMH volume. While the correlation was significant in the FWM (*r* = −0.360; *p* = 0.047, Figure 2A, Table 2), a trend toward significance was observed in the PCC (*r* = −0.353; *p* = 0.052; Figure 2B, Table 2).

**Figure 2.**
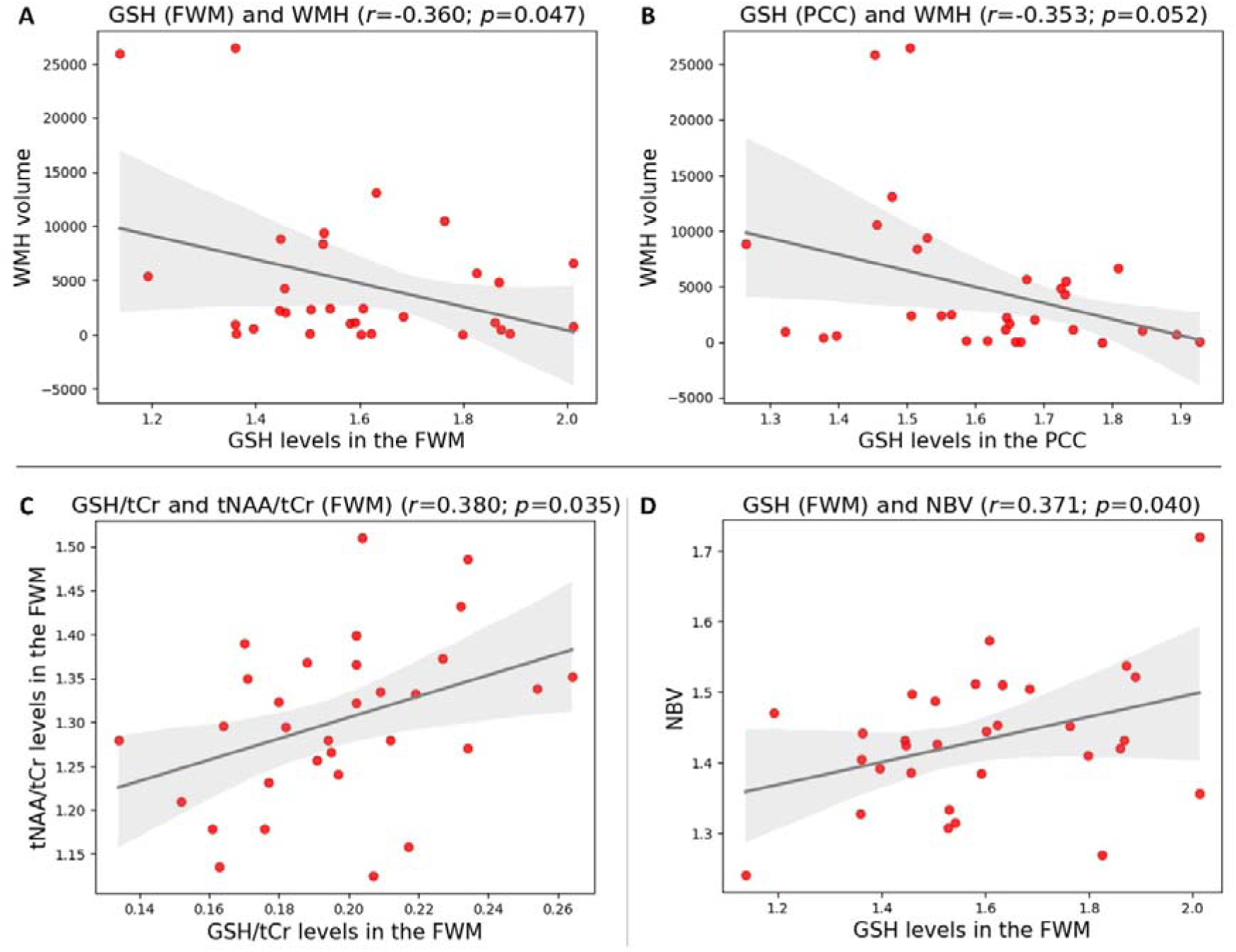
Scatter plots showing associations between **A)** GSH levels in the FWM and WMH volume (significant association), **B)** GSH levels in the PCC and WMH volume (trend toward significant association), **C)** GSH/tCr and tNAA/tCr levels in the FWM (significant association), and **D)** GSH levels in the FWM and NBV (significant association). The black line represents a linear fit to the data, and the grey area reflects 95% confidence interval. Pearson’s *r* and corresponding *p*-values are provided. *Abbreviations*: FWM, frontal white matter; GSH, glutathione; NBV, normalized brain volume; tCr, total creatine; PCC, posterior cingulate cortex; tNAA, total N-acetyl aspartate; WMH, white matter hyperintensity.

**Table 2.** Pearson correlations between GSH levels and other variables of interest. *Abbreviations:* FWM, frontal white matter; GSH, glutathione; NBV, normalized brain volume; nHCPv, normalized hippocampal volume; tCr, total creatine; PCC, posterior cingulate cortex; tNAA, total N-acetyl aspartate; WMH, white matter hyperintensity.

|  | Region | WMH | tNAA | NBV | nHCPv | Age |
| --- | --- | --- | --- | --- | --- | --- |
| GSH | PCC | $r = -0.353$<br>$p = 0.052$ | $r = 0.157$<br>$p = 0.399$ | $r = 0.212$<br>$p = 0.253$ | $r = 0.330$<br>$p = 0.070$ | $r = -0.469$<br>$p = 0.008$ |
| | FWM | $r = -0.360$<br>$p = 0.047$ | $r = 0.259$<br>$p = 0.160$ | $r = 0.371$<br>$p = 0.040$ | $r = 0.086$<br>$p = 0.646$ | $r = -0.180$<br>$p = 0.332$ |

### 3.3 Association between metabolite markers of oxidative stress and neuroaxonal integrity

We found a positive significant association between GSH/tCr and tNAA/tCr levels in the FWM (*r* = 0.380; *p* = 0.035; Figure 2C, Table 3).

**Table 3.** Pearson correlations between GSH/tCr levels and other variables of interest. *Abbreviations:* FWM, frontal white matter; GSH, glutathione; NBV, normalized brain volume; nHCPv, normalized hippocampal volume; tCr, total creatine; PCC, posterior cingulate cortex; tNAA, total N-acetyl aspartate; WMH, white matter hyperintensity.

|  | Region | WMH | tNAA/tCr | NBV | nHCPv | Age |
| --- | --- | --- | --- | --- | --- | --- |
| GSH/tCr | PCC | $r = -0.199$<br>$p = 0.283$ | $r = 0.035$<br>$p = 0.850$ | $r = -0.017$<br>$p = 0.927$ | $r = 0.105$<br>$p = 0.573$ | $r = -0.305$<br>$p = 0.095$ |
| | FWM | $r = -0.140$<br>$p = 0.453$ | **$r = 0.380$**<br>**$p = 0.035$** | $r = 0.330$<br>$p = 0.070$ | $r = 0.002$<br>$p = 0.993$ | $r = -0.091$<br>$p = 0.625$ |

### 3.4 Association between oxidative stress markers and brain tissue integrity

We found GSH levels in the FWM to be significantly associated with normalized brain volume (*r* = 0.371; *p* = 0.040; Figure 2D, Table 2), but not hippocampal volume. GSH levels in the PCC were not associated with normalized brain volume or hippocampal volume (Table 2).

### 3.5 Metabolite levels in the FWM and PCC

In the FWM compared to the PCC, we found (a) GSH/tCr level to be significantly higher, while (b) tNAA/tCr and tNAA levels to be significantly lower (*p* < 0.001) (Figure 3, Table 1). No significant differences were found for GSH levels between the FWM and the PCC (Table 1).

**Figure 3.**
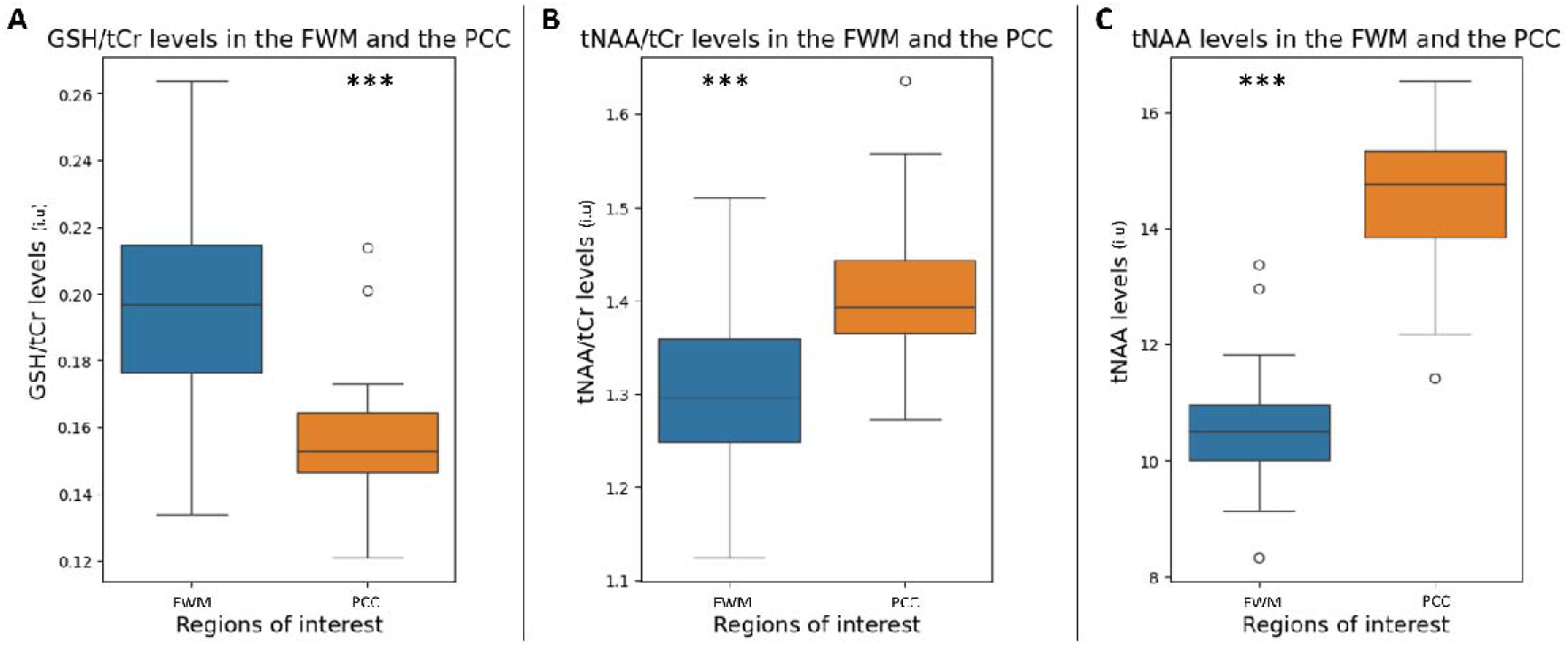
Boxplots of **A)** GSH/tCr levels in the FWM (left) and the PCC (right), **B)** tNAA/tCr levels in the FWM (left) and the PCC (right), and **C)** tNAA levels in the FWM (left) and the PCC (right). *** denotes *p* < 0.001. *Abbreviations*: FWM, frontal white matter; GSH, glutathione; IU, institutional units; tCr, total creatine; PCC, posterior cingulate cortex; tNAA, total N-acetyl aspartate; WMH, white matter hyperintensity.

### 3.6 Association between cognition and other markers

MoCA scores were not significantly associated with any markers.

## 4. DISCUSSION

Studies have shown that MRS is sensitive to metabolic alterations in the AD continuum. In this study, we explored the relationship between MRS-detected brain GSH levels, a marker of oxidative stress, and both vascular and neuronal injury markers, as well as overall brain integrity and cognition in MCI individuals. By probing these associations, we offer new insights into how oxidative stress may contribute to neurovascular injury, highlighting GSH’s potential role in linking vascular and neuronal pathologies in early-stage AD.

### 4.1 Oxidative stress as a driver of vascular injury in AD

Our study revealed a significant inverse relationship between MRS-detected brain GSH levels and WMH volume in MCI participants. This suggests that increased oxidative stress during the early stages of AD may drive or be driven by vascular injury, shedding light on an important but under-researched link between GSH, oxidative stress, and vascular pathology in AD. This finding aligns with observations in non-AD conditions, such as COVID-19 survivors [30], where decreased GSH levels were associated with higher WMH volume, suggesting that reduced GSH levels contribute to white matter damage and increased WMH burden [30]. Further supporting our results, studies have linked lower plasma levels of nitric oxide and α-tocopherol (vitamin E) to more severe WMH burdens in middle-aged to older adults [45,46]. Both these antioxidants are known to interact with GSH [17].

Our observed relationship between brain GSH and WMH volume is particularly relevant when considering the pathogenic mechanisms that contribute to WMHs, such as brain endothelial dysfunction and blood-brain barrier leakage [47–49]. Both pathological processes have been documented in MCI individuals [50–54] and mouse models of AD [55–57], with oxidative stress identified as a key contributor [50,53]. GSH not only protects brain endothelial cells from oxidative stress but also supports their repair and proliferation following injury [58,59]. In the context of blood-brain barrier damage, GSH delays the transition of endothelial cells from DNA replication to cell division, allowing additional time for DNA repair [60]. Additionally, GSH helps maintain blood-brain barrier integrity by preserving tight junction proteins like claudin 5 [59].

### 4.2 GSH and neuronal brain injury markers

Our findings revealed significant positive correlations in the FWM between (a) GSH/tCr and tNAA/tCr ratios – with NAA considered a key neuroaxonal marker [61], and (b) GSH levels and normalized brain volume, a marker of brain tissue preservation. These results highlight increased oxidative stress as an important contributor to increased neuronal injury and subsequent brain atrophy – a key feature of MCI and AD dementia [4]. Although human studies on the relationship between GSH and neuronal health in the AD continuum are scarce, our findings align with (a) increased death in neurons cultured from AD transgenic mice following GSH depletion [62], and (b) neuronal injury/loss in mice lacking a GSH synthesis enzyme [63,64].

In contrast to GSH, NAA levels have been better investigated in AD. Lower NAA levels (or its ratio) have been observed in MCI [65] and AD individuals across several brain regions, including the posterior cingulate gyrus, hippocampus, and frontoparietal cortex [26,66,67]. Additionally, (a) reduced NAA levels have been associated with an increased risk of conversion from MCI to AD dementia [65,68], and (b) lower NAA/tCr early in the AD continuum were associated with amyloid-beta and tau – two hallmark AD pathologies [69].

### 4.3 GSH and cognition

We found no significant association between GSH levels in PCC or FWM and cognition, as assessed using MoCA. Notably, no studies have specifically examined the relationship between GSH levels and cognition using MoCA within the AD continuum, though other cognitive tests have explored this connection. Studies comparing GSH levels in the AD continuum to cognitively unimpaired individuals have reported the following associations: Lower hippocampal GSH levels in MCI, detected via 3T MRS, were significantly associated with worse global cognition on the Mini Mental State Examination (MMSE) [22]. Similarly, lower GSH levels in the postmortem frontal cortex in MCI correlated with poorer premortem MMSE scores [25]. Additionally, lower 3T MRS-detected GSH levels in both the hippocampus and frontal cortex in AD were linked to declines in both global cognition (MMSE, Clinical Dementia Rating) and executive functions (Trail Making Test A and B) [22]. Studies along the AD continuum have also assessed GSH in blood, revealing lower levels than in cognitively unimpaired individuals. In whole blood and plasma, lower GSH levels in MCI and AD were associated with worse cognition as assessed by the MMSE [70].

Commensurate with the identification of aging as the most significant risk factor for AD, a longitudinal study in older healthy adults (65+ years) found that higher baseline plasma GSH levels were associated with lower AD risk and better cognitive performance, in particular executive function [29]. In contrast, two cross-sectional studies in the same age group reported no association between MRS-detected brain GSH levels (in the medial frontal, sensorimotor, and occipital regions) and cognition as measured by MoCA [18].

### 4.4 Antioxidant therapies as potential disease-modifying treatments in AD

Given the role of oxidative stress and GSH in AD pathogenesis and progression, including cognitive decline, several clinical trials are testing GSH and associated compounds as disease-modifying treatments [15,18]. Clinical trials have shown that it is possible to increase GSH levels in the brain following dietary supplement intake [18]. Specifically, supplementation with N-acetyl cysteine (NAC) has been shown to (a) increase GSH levels by providing cysteine, the rate limiting compound in GSH synthesis [71], (b) protect cells from amyloid-beta-induced apoptosis [71], and (c) improve cognition in the senescence-accelerated mouse-prone 8 (SAMP8) model overexpressing the amyloid precursor protein [71] and in human psychosis [72]. Antioxidant therapy with catalase in mice overexpressing the amyloid precursor protein has been shown to completely reverse cerebrovascular dysfunction [55,73], with catalase being one of the two systems, together with the GSH system, for detoxifying hydrogen peroxide, a major reactive oxygen species [74]. In healthy middle-aged and older humans, antioxidant-promoting dietary intake, such as dairy food or green tea, has been shown to (a) increase brain GSH levels, and (b) increase superoxide dismutase and glutathione peroxidase and decrease malondialdehyde levels in serum, respectively [18,75], resulting in improved cognition as assessed by MoCA, Hopkins Verbal Learning Test (HVLT), TMT-B, and Victoria Stroop test interference scores [75]. Together with improved cognition and altered oxidative stress markers, green tea consumption was also shown to alter AD markers such as serum pTau_181_, amyloid-beta_42_, and total amyloid-beta levels [75], showcasing the potential relevance of improving antioxidant capacity in the management of AD.

### 4.5 Limitations

The main limitation of our study was our small sample size which may have influenced our ability to detect strong significant associations. Therefore, this work constituted a preliminary study providing a strong background and novel findings to be validated and reproduced in our ongoing work using a bigger cohort and improved spectral resolution.

## 5. CONCLUSION

Our study sheds light on the relationship between oxidative stress, vascular and brain injury markers, and cognitive function in individuals with MCI. Moving forward, longitudinal studies with comprehensive cognitive assessments and larger cohorts are warranted to validate our findings and further elucidate the causal relationships between oxidative stress, brain injury markers, and cognitive decline in MCI.

## Data Availability

All data produced in the present work are contained in the manuscript

## ACKNOWLEDGMENTS

The authors thank Annie Karabadjian, MBA, BSc, Medscope Inc. for her contributions in preparing most of the study documents for the parent MCI study (NCT03448055) from which the baseline data were analyzed for the current manuscript. We would also like to thank Dr. Ali Filali-Mouhim, PhD, from the Centre de Recherche de l’Institut Universitaire de Gériatrie de Montréal (CRIUGM) for his input for the statistical analysis. We also thank our intern, Ms. Olohita Ayodele from Dawson College, Montreal, Canada, for her assistance in data extraction.

## CONSENT STATEMENT

The study was approved by the research ethics boards at the CIUSSS-Centre-Ouest de Montreal (Jewish General Hospital) and the McGill University Health Centre (Montreal Neurological Institute-Hospital), and informed consent was obtained for all participants.

