## Supplementary Table S1 for "Investigating the oxidative stress-vascular brain injury axis in mild cognitive impairment of the Alzheimer type"

**Supplementary Table S1: MRS in MRS checklist** [1]

**1. Hardware**

| Field strength (tesla, T) | 3 T |
| --- | --- |
| Manufacturer | Siemens |
| Model (software version) | Prisma (VE11C) |
| Radio-frequency coils: nuclei (transmit/receive), number of channels, type, body part | 64 channel phased array receive head coil |
| Additional hardware | N/A |

**2. Acquisition**

| Pulse sequence | SPin Echo full Intensity Acquired Localized (SPECIAL) |
| --- | --- |
| Volume of interest (VOI) location | **Posterior cingulate cortex (PCC)**: oblique axial voxel parallel to the line connecting the inferior aspect of the genu and splenium of the corpus callosum (callosal line). Voxel was positioned on the mid-line grey matter posterior to the splenium of the corpus callosum to intersect PCC and precuneus  **Left frontal white matter (FWM)**: superior to the lateral ventricles and corpus callosum, subjacent to the cerebral cortex |
| Nominal voxel size | PCC: 30 mm x 30 mm x 20 mm  FWM: 15 mm x 30 mm x 15 mm |
| Repetition time/Echo time (TR/TE) [ms] | 3000/8.5 |
| Total number of excitations or acquisitions per spectrum | Water suppressed (PCC): 128 averages  Water suppressed (FWM): 256 averages  Non-water-suppressed: 16 averages |
| In time series for kinetic studies  i. Number of averaged spectra (N/A) per time point  ii. Averaging method  iii. Total number of spectra (acquired/in time series) | N/A |
| Additional sequence parameters (spectral width [Hz], number of spectral points, frequency offset), 2D field of view (FOV), matrix size, acceleration factors, sampling method | Spectral width: 2000 Hz  Number of spectral points: 4096  Frequency offset: -2.3 ppm  Acceleration: None  Sampling method: Conventional |
| Water suppression method | VAriable Power and Optimized Relaxation delays (VAPOR) |
| Shimming method | Siemens “Brain” B0 shim mode |
| Triggering or motion correction method | N/A |

**3. Data analysis methods and outputs**

| Analysis software | FID-A for data processing and LCModel for metabolite quantification |
| --- | --- |
| Processing steps deviating from quoted reference or product | N/A |
| Output measure | LCModel metabolite concentrations (N-acetylaspartate [NAA] and glutathione [GSH]) and ratios to creatine (Cr) |
| Quantification references and assumptions, fitting model assumptions | For LCModel, an internal unsuppressed water reference was used for metabolite quantification. The unsuppressed water scan was also used for eddy current correction |

**4. Data quality**

| Reported variables (signal-to-noise ratio [SNR], linewidth [with reference peaks]) | Metabolite concentrations (in institutional units)  Metabolite ratios to Cr |
| --- | --- |
| Data exclusion criteria | SNR < 20; linewidth (LW) > 10 Hz No data was excluded as all datasets passed quality check |
| Quality measures of post processing model fitting (e.g., Cramer-Rao lower bound [CRLB], goodness of fit, standard deviation of residual) | SNR, LW, goodness of fit CRLB ranges for NAA: 0-5%  CRLB ranges for GSH: 0-8% |
| Sample spectrum | Figure 1 |
